# Colopathy Associated with Pentosan Polysulfate Use

**DOI:** 10.1101/2023.04.03.23288071

**Authors:** Emily H. Jung, Wei Zheng, Ryan J. Weiss, Namita E. Mathew, Benjamin I. Meyer, Azhar Nizam, Heba Iskandar, Nieraj Jain

## Abstract

**Introduction:** We describe a novel colopathy associated with pentosan polysulfate (PPS) use and measure the strength of the drug-disease association.

**Methods:** Two-part investigation. In the cohort study of individuals with a history of prior long-term PPS use, case histories were obtained and gastrointestinal disease course was followed with review of endoscopy records and histopathology specimens. Findings were summarized with descriptive statistics. In the cross-sectional study of individuals with interstitial cystitis, drug exposure and medical histories were obtained for patients seen at a single clinical center. Strength of association between PPS use and diagnoses of inflammatory bowel disease (IBD) and/or irritable bowel syndrome (IBS) was measured with multivariate logistic regression.

**Results:** In the cohort study of 13 participants, median PPS exposure was 2.04 kg (0.99–2.54). Eleven (84.6%) developed symptoms suggestive of IBD and/or IBS after initiation of PPS therapy. Of the 10 participants whose endoscopic and histopathologic findings we reviewed, six had abnormal-appearing colonic mucosa on endoscopy and all 10 had abnormal mucosal changes on histology. Clinical and histologic improvement was observed after PPS cessation. In the cross-sectional study of 219 subjects with interstitial cystitis, PPS use was a statistically significant predictor of both the IBD [adjusted odds ratio=3.3 (95% confidence interval, 1.2–8.8, p=0.02)] and the composite IBD+IBS [adjusted odds ratio=3.3 (95% confidence interval, 1.5–7.3, p=0.002)] outcomes.

**Discussion:** We describe a strong association between PPS use and a clinical diagnosis of IBD and/or IBS. Histopathologic findings suggest a novel drug-associated colopathy, with some subjects requiring colectomy for dysplasia.

---

Interstitial cystitis (IC), also known as bladder pain syndrome, is a chronic condition characterized by suprapubic and pelvic pain as well as lower urinary tract symptoms. It affects more than one million U.S. adults, predominantly women.^1^ Although the pathophysiology of IC remains unclear, abnormalities in the bladder uroepithelium may expose the bladder wall to irritants within the urine.^2^ Under the 2022 American Urological Association guidelines, there are four recommended oral medications for the treatment of IC.^1^ Of these, pentosan polysulfate sodium (PPS) [ELMIRON, Janssen Pharmaceuticals, Titusville, NJ] is the only oral drug approved by the U.S. Food and Drug Administration (FDA) for management of IC.^3^

Pentosan polysulfate is a semi-synthetic sulfated polysaccharide that is structurally related to the cellular glycosaminoglycan heparan sulfate. It is postulated that PPS may integrate into and support the inner lining of the bladder urothelium.^4^ PPS received FDA approval for IC management in 1996 and has been associated with a number of adverse events, with diarrhea (3.9%), alopecia (3.9%), and nausea (3.7%) being the most frequently reported in the original FDA medical review documents.^5^ Five of the 33 serious adverse drug reactions were gastrointestinal (GI) in nature, and attributed to an osmotic load effect induced by undigested or unabsorbed PPS. Notably, the FDA medical review noted that the disease IC itself is associated with an increased incidence of inflammatory bowel disease (IBD).^5,6^ Indeed, studies show that approximately 2% of patients with classic IC have IBD, compared to a 0.07% prevalence in the general population.^7,8^

In 2018, our group described a novel vision-threatening retinal disease associated with long-term PPS use, which subsequently led to a drug labeling change.^9,10^ During our ongoing investigations of PPS maculopathy, we identified a high prevalence of IBD, with many patients reporting “ confusing” or “ atypical” diagnoses from prior GI evaluations. Intriguingly, dextran sodium sulfate (DSS), a structurally related semi-synthetic sulfated polysaccharide, is routinely used to artificially induce acute and chronic colitis in animal models (Figure 1).^11,12^ We hypothesize that the GI diseases observed in our PPS-exposed patients may represent a drug toxicity. Of note, a recent retrospective series identified a high rate of colonic dysplasia in patients with IBD who had used PPS.^13^ Herein, we report our explorations into a novel association between PPS use and incident colopathy with a cohort study of participants with long-term PPS exposure and a cross-sectional study of individuals with IC.

**Figure 1.**
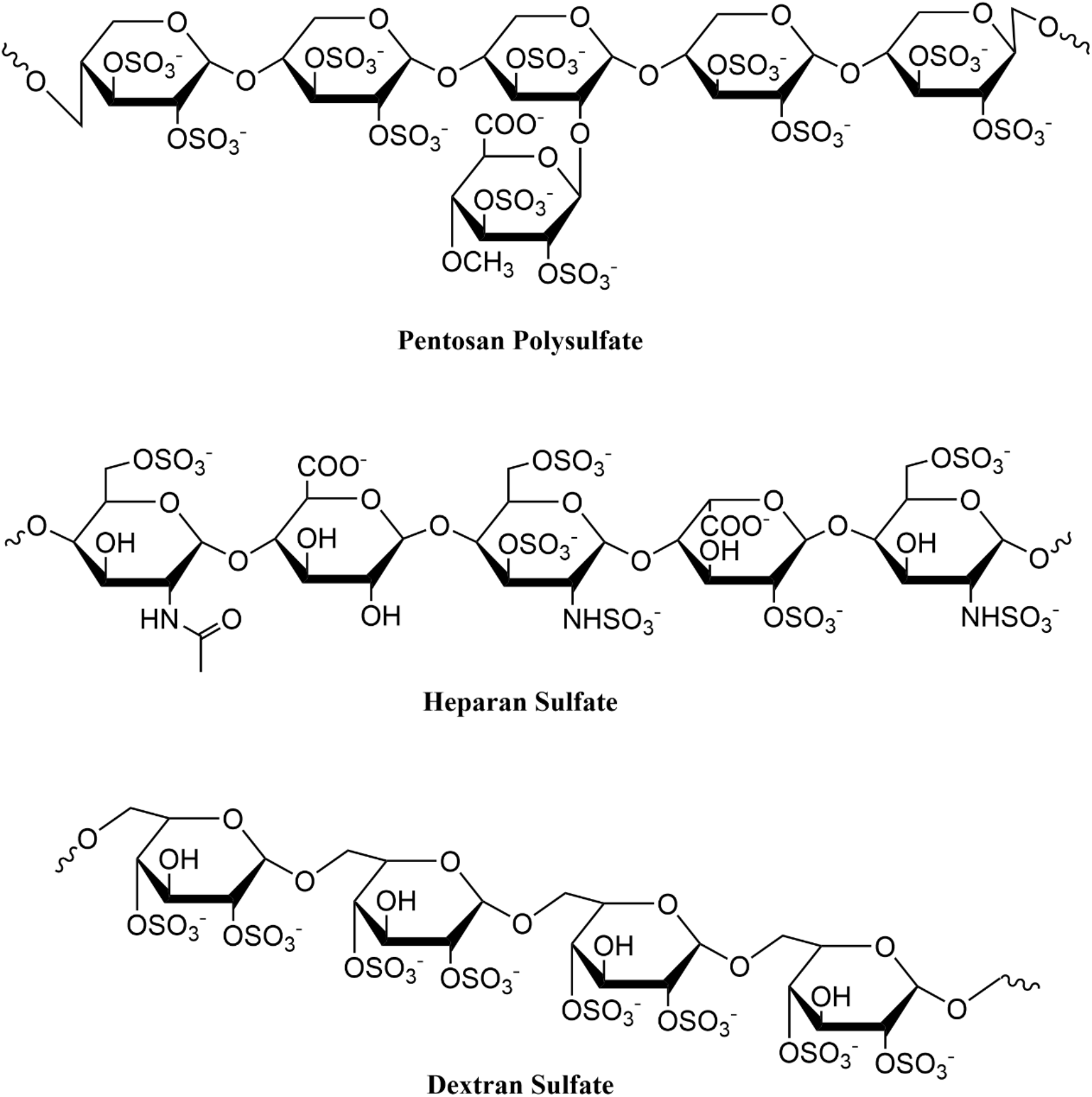
Chemical structures of pentosan polysulfate, heparan sulfate, and dextran sulfate.

## Methods

The study was conducted at Emory University (Atlanta, GA) and received approval from the Emory University Institutional Review Board. The study followed the tenets set forth by the Declaration of Helsinki.

### Part I: Cohort Study

Fourteen participants with PPS maculopathy enrolled in a four-year prospective study of their ocular and systemic health, prior to our concerns regarding an association between PPS and colopathy. The study protocol was subsequently amended to permit more detailed investigations into systemic health. All participants had discontinued PPS use prior to the baseline visit. Study enrollment occurred between 2019 and 2020. All participants provided written informed consent and Health Insurance Portability and Accountability Act authorization. In addition to ophthalmic investigations, medical records pertaining to GI symptoms and diagnoses, endoscopy findings, and other medical history were reviewed. Participants were queried for additional medical history and to verify content from the medical records. Case histories were reviewed by a gastroenterologist with expertise in inflammatory bowel disease (HI). Outside endoscopy reports were reviewed and biopsy specimens were retrieved for further evaluation by an expert gastrointestinal pathologist (WZ). Demographic and clinical characteristics were summarized using descriptive statistics.

### Part II: Cross-Sectional Study

To further explore the strength of association between PPS and gastrointestinal disease, a cross-sectional study was conducted among individuals with a history of interstitial cystitis seen at our facility. The requirement for informed consent for this portion of the study was waived. Local billing records, electronic medical records, and pharmacy databases were queried for patients with a diagnosis of IC seen at the Emory Eye Center between May 2014 and October 2018. For each patient with IC, all available Emory Healthcare medical records were reviewed to identify those with a diagnosis of IBD and/or irritable bowel syndrome (IBS). Records were also evaluated for exposure to other IC therapies and medications associated with the development or exacerbation of IC.

Odds ratios (OR) were computed to quantify the association between each drug exposure and two separate outcomes based on clinical diagnoses: IBD, and a composite outcome of IBD and/or IBS (IBD+IBS). Crude (unadjusted) associations were assessed as follows: two-sample t-tests were used to compare the average age for participants with and without the outcomes, and either chi-square tests or, where appropriate, Fisher’s exact tests were used for associations between categorical covariates and the outcomes. Multivariable logistic regression was performed for both outcomes, with PPS use, age, gender, smoking status, and nonsteroidal anti-inflammatory drug (NSAID) use as covariates. Adjusted ORs were estimated and p-values for type III tests determined. Significance level was set at p<0.05 (confidence level 95%). Statistical analysis was performed using SAS version 9.4 (SAS Institute, Cary, NC).

## Results

### Cohort Study

Fourteen individuals with PPS maculopathy were enrolled in a prospective study of their ocular and systemic health (Table 1). One individual withdrew following the baseline visit and was excluded from all analyses. All 13 included participants had a longstanding diagnosis of IC previously managed with PPS and all experienced gastrointestinal symptoms while on PPS. Twelve (92.3%) were female and twelve (92.3%) were White. Median (interquartile range [IQR]) age at study enrollment was 62 years (48–65). Median cumulative PPS exposure was 2.04 kg (0.99–2.54) over a median duration of 14 years (9–18).

**Table 1.** Demographics and PPS exposure history by study subject (cohort study).

| **Subject ID** | **Age/Sex/Race** | **Body Mass Index (kg/m<sup>2</sup>)** | **Year of IC diagnosis** | **Duration of PPS Intake (years)** | **Mean PPS Daily Dose (mg)** | **Cumulative PPS Exposure (kg)** | **Smoking History** | **NSAID Use** |
| --- | --- | --- | --- | --- | --- | --- | --- | --- |
| 1 | 30s/F/W | 38.1 | 2003 | 14 | 400 | 2.04 | Never |  |
| 2 | 60s/F/W | 26.2 | 2011 | 7.4 | 300 | .84 | Never |  |
| 3 | 60s/F/W | 27.8 | 1994 | 19.9 | 300 | 2.18 | Never |  |
| 4 | 60s /F/W | 29.0 | 1998 | 17 | 400 | 2.48 | Never | Y (Daily use for >10 years) |
| 5 | 40s/F/W | 26.2 | 2014 | 3 | 400 | .44 | Never |  |
| 6 | 40s/F/W | 35.4 | 2003 | 14 | 400 | 2.04 | Never |  |
| 7 | 60s/F/W | 20.4 | 1978 | 12 | 300 | 1.31 | Never |  |
| 8 | 50s/F/W | 21.1 | 1995 | 18.3 | 400 | 2.68 | Never |  |
| 9 | 30s/F/AA | 22.2 | 1999 | 9 | 300 | .99 | Never |  |
| 10 | 70s/F/W | 19.3 | 1995 | 21.8 | 400 | 3.19 | Never |  |
| 11 | 70s/M/W | 26.7 | 2000 | 19.8 | 400 | 2.85 | 7-pack-year; quit in his 20s |  |
| 12 | 50s/F/W | 25.2 | 2007 | 6.4 | 300 | .70 | Never |  |
| 13 | 60s/F/W | 25.1 | 2002 | 17.4 | 400 | 2.54 | Never |  |

Eleven (84.6%) participants experienced the onset of symptoms suggestive of IBD or IBS following the initiation of PPS therapy (Table 2). Median time between PPS therapy initiation and symptom onset was 4.9 years (3.5–6.2). Initial symptoms included diarrhea (n=7), abdominal pain (n=2), stool with mucus and blood (n=2), tenesmus (n=1), fecal urgency (n=1), and fecal incontinence (n=1). For the two participants with symptom onset prior to PPS use, one (subject 1) was diagnosed with lactose intolerance as a child and one (subject 6) had received a prior diagnosis of IBS.

**Table 2.** Gastrointestinal history and disease course by study subject (cohort study).

| Subject ID | Duration between PPS initiation and initial GI symptoms (years) | GI Symptoms |  |  |  |  |  |  |  | Duration between PPS initiation and earliest abnormal histopathologic findings (years) | GI Diagnoses |  | GI surgical history | IBD Medications |
| --- | --- | --- | --- | --- | --- | --- | --- | --- | --- | --- | --- | --- | --- | --- |
|  |  | Abdominal pain | Constipation | Diarrhea | Incontinence | Mucus in stool | Hematochezia | Tenesmus | Urgency |  | IBD | IBS |  |  |
| 1 | -1.3 |  |  |  |  | x* |  |  |  | - |  |  | Abdominal lipoma tumor removal |  |
| 2 | 4.3 |  | x* | x |  |  |  |  |  | 4.9 | UC, MC |  | Colectomy (colitis-associated dysplasia) | Mesalamine (discontinued) |
| 3 | 7.8 | x |  |  |  |  | x | x | x | 8.2 | CD |  | Cholecystectomy | Balsalazide |
| 4 | 5.3 |  |  | x |  |  |  |  |  | 17.3 | CD | x | Cholecystectomy, adhesiolysis | Budesonide |
| 5 | 0.8 |  | x<sup>#</sup> | x |  |  |  |  |  | 3.0 | Indeterminate |  | Appendectomy, sigmoid colectomy (fixed loop of sigmoid colon in the pelvis following hysterectomy) |  |
| 6 | -4.4 | x* | x* |  |  |  |  | x* |  | - |  | x |  |  |
| 7 | 0.8 |  |  | x |  |  |  |  |  | 1.5 | UC |  |  |  |
| 8 | 7.0 | x |  | x |  |  | x |  |  | 7.0 | UC |  |  | Mesalamine (discontinued) |
| 9 | 3.0 |  |  |  |  |  |  |  |  | - |  | x | Appendectomy |  |
| 10 | 4.0 |  |  | x |  |  |  |  |  | 21.5 |  | x | Cholecystectomy |  |
| 11 | 4.9 |  |  | x |  |  |  |  |  | 15.7 | UC |  | Colectomy (colitis-associated dysplasia) | Mesalamine (discontinued) |
| 12 | 5.0 |  |  | x |  |  |  |  |  | 5.9 | UC |  |  | Mesalamine |
| 13 | 9.9 |  |  |  | x |  |  |  |  | 11.4 | UC |  |  | Mesalamine |

\* = symptoms occurred prior to starting PPS

All 11 participants with symptom onset after starting PPS underwent at least one colonoscopy while on the drug, and all 11 received a clinical diagnosis of IBD and/or IBS. Nine participants received a diagnosis of IBD, including five with ulcerative colitis (UC), two with Crohn’s disease, one with microscopic colitis, and one with indeterminate colitis. Three of the 11 participants were diagnosed with IBS, one of whom also had an IBD diagnosis.

Endoscopy reports, pathology reports, and histology slides were reviewed for 10 of these 11 participants (Table 3). On endoscopy, three (27.3%) of the 11 had both mucosal ulcerations and polyps, one (9.1%) had mucosal ulcerations only, two had polyps only (18.2%), and four (36.4%) had normal-appearing colonic mucosa. Histology showed abnormal mucosal changes in all 10 participants whose reports and slides we reviewed. Median time from PPS initiation to the earliest abnormal histopathologic finding was 7.6 years (5.2–14.6). Seven (63.6%) participants exhibited chronic mucosal injury and five (45.5%) had histologic findings of focal minimal to mild active colitis. Three (27.3%) had only chronic mucosal injury with no significant lamina propria inflammation and two (18.2%) had only reactive changes. Specific findings included crypt architectural distortion (n=8), patchy lamina propria fibrosis and thickening of the muscularis mucosae (n=4), patchy lamina propria lymphoplasmacytic inflammation with increased eosinophils (n=5), increased crypt epithelial apoptotic bodies (n=4), fibrin-appearing eosinophilic amorphous deposition in lamina propria capillaries and/or stromal cells (n=2), foamy cytoplasm (n=2), superficial granulomas (n=1), and low-grade dysplasia (n=1) (Table 3; Figure 2).

**Table 3.** Endoscopy and histology findings by study subject (cohort study).

| Subject ID | Year of PPS initiation | Year of PPS cessation | Year and indication for colonoscopy | Endoscopy Findings |  |  |  | Histology Findings |  |  |  |
| --- | --- | --- | --- | --- | --- | --- | --- | --- | --- | --- | --- |
|  |  |  |  | Polyps | Mucosal erythema/erosions | Aphthous ulcerations | Macroscopic colitis-appearing mucosa | Crypt distortion/dropout/branching | Eosinophils (E) and neutrophils (N) | Thickening of the muscularis mucosae | Other findings and general histopathologic comments |
| 1 | 2004 | 2018 | - | - | - | - | - | - | - | - | - |
| 2 | 2011 | 2019 | 2005 Constipation | 1 | 0 | 0 | 0 | n/a (only polyps biopsied) | n/a | n/a |  |
|  |  |  | **2016 Diarrhea** | **1** | **0** | **0** | **0** | **1** | **0** | **0** | **Mild chronic mucosal injury with no significant inflammation.** |
|  |  |  | 2019 UC/MC history, screening/asymptomatic | 1 | 0 | 0 | 0 | 0 | 0 | 0 | Benign colonic mucosa demonstrating no significant histopathologic change. |
|  |  |  | *2020 Screening/asymptomatic | 1 | 0 | 0 | 0 | n/a (only polyps biopsied) | n/a | n/a |  |
|  |  |  | *2021 Screening/asymptomatic | 1 | 0 | 0 | 0 | n/a (only polyps biopsied) | n/a | n/a |  |
| 3 | 1998 | 2018 | **2006 Hematochezia** | **0** | **1** | **1** | **1** | **1** | **N** | **1** | **Acute and chronic focally crypt-destructive colitis (lymphoplasmacytic inflammation). Mildly increased crypt epithelial apoptotic bodies. Definitive granulomata are not identified.** |
|  |  |  | **2007 CD history** | **0** | **1** | **1** | **1** | **1** | **E** | **0** | **Epithelium is slightly hypermucinous. Inflammatory cell population within the lamina propria is not increased, encroaching, or shifted toward acuity. Minimal crypt architectural changes and focal mild active chronic inflammation.** |
|  |  |  | **2010**<br>**CD history** | **0** | **n/a** | **n/a** | **n/a** | **1** | **N** | **0** | **Hypermucinous crypts. One fragment with fairly prominent lymphoid nodule with a germinal center. Mild active chronic inflammation and architectural disorder, nonspecific.** |
|  |  |  | 2021<br>CD history | 1 | 0 | 0 | 0 | 0 | 0 | 0 | Colonic mucosa with melanosis coli; otherwise no significant histopathology. No evidence of active, chronic, or microscopic colitis.<br>No granulomas identified. |
|  |  |  | 2021<br>Earlier in the year, performed piecemeal removal of adenoma | 1 | 0 | 0 | 0 | 1 | 0 | 0 | Colonic mucosal villous change suggestive of chronic mucosal injury. |
|  |  |  | 2022<br>CD history | 1 | 0 | 0 | 0 | 1 | 0 | 0 | No granulomas are identified, no increase in inflammation. Colonic mucosa with patchy mild architectural irregularity/regenerative features, suggesting healing from prior injury. |
| 4 | 2001 | 2018 | *2018<br>Diarrhea | 0 | 0 | 0 | 0 | 1 | E | 1 | Chronic colonic mucosal injury characterized by crypt dropout, architectural disarray. Patchy lamina propria fibrosis and thickening of the muscularis mucosae. Mildly increased crypt epithelial apoptotic bodies. Patchy increased lamina propria eosinophils. Small nonspecific superficial granulomas in the proximal colon. |
| 5 | 2012 | 2015 | **2012**<br>**Specimen**<br>**obtained from**<br>**sigmoidectomy** | **0** | **0** | **0** | **0** | **0** | **0** | **0** | **Reactive colonic mucosa with occasional small lymphoid aggregates. Ganglion cells present in the intermyenteric Auerbach's plexus.** |
|  |  |  | **\*2015**<br>**Diarrhea** | **0** | **0** | **0** | **0** | **1** | **E** | **1** | **Chronic colonic mucosal injury characterized by crypt dropout, architectural disarray. Patchy lamina propria fibrosis and thickening of the muscularis mucosae. Mildly increased crypt epithelial apoptotic bodies. Reactive ileal mucosa.** |
| 6 | 2004 | 2018 | - | - | - | - | - | - | - | - | - |
| 7 | 2006 | 2018 | **2007**<br>**Diarrhea** | **0** | **1** | **1** | **1** | **1** | **E,N** | **0** | **Histologic features in all three biopsies are similar with the sigmoid colon more** pronounced. Edema is present within the lamina propria of all three specimens as well as some reactive architectural glandular changes. Sigmoid colon biopsy shows a moderate increase in inflammatory cells in the lamina propria consisting of lymphocytes, plasma cells, eosinophils, and neutrophils. |
| 8 | 2001 | 2019 | 2008 Diarrhea and hematochezia | n/a | n/a | n/a | 0 | n/a | n/a | n/a | No procedure/pathology reports available. GI clinic note states no evidence of active disease was noted on colonoscopy. |
|  |  |  | *2013 UC history | 0 | 0 | 0 | 0 | 0 | 0 | 0 | Fibrin-appearing eosinophilic amorphous deposition in lamina propria capillaries and/or some stromal cells. Melanosis coli. Focally increased reactive histiocytes in the lamina propria. |
|  |  |  | *2019 Screening/asymptomatic | 0 | 0 | 0 | 0 | 0 | 0 | 0 | Multiple mucosal lymphoid aggregates and occasional foamy cytoplasm of stromal and epithelial cells. No inclusion bodies/globules. |
|  |  |  | *2019 Screening/asymptomatic | 0 | 0 | 0 | 0 | 0 | 0 | 0 | No lymphoid aggregates, no foamy cytoplasm, and no inclusion bodies/globules. |
| 9 | 1999 | 2008 | - | - | - | - | - | - | - | - | - |
| 10 | 1996 | 2017 | *2017 Diarrhea | 0 | 0 | 0 | 0 | 0 | E,N | 0 | Focal lamina propria lymphoplasmacytic inflammation with an increase in eosinophils, lymphoid aggregates, and focal red blood cell extravasation. |
| 11 | 2000 | 2019 | 2012 Diarrhea | n/a | n/a | n/a | 0 | n/a | n/a | n/a | No procedure/pathology reports available. GI clinic note states no evidence of active disease was noted on colonoscopy. |
|  |  |  | *2015 Diarrhea | 1 | n/a | n/a | n/a | 1 | N | 0 | Low grade dysplasia arising in a background of chronic mucosal injury. Foamy cytoplasm noted |
|  |  |  | *2016 UC history | 1 | n/a | n/a | n/a | 1 | E,N | 0 | Crypt architectural distortion. Patchy increased lamina propria lymphoplasmacytic inflammation with an increase in eosinophils and scattered neutrophils. Mildly increased crypt epithelial apoptotic bodies. Indefinite for dysplasia in the inflamed areas. |
| 12 | 2008 | 2014 | **\*2013**<br>**Diarrhea** | **0** | **1** | **1** | **1** | **1** | **E,N** | **0** | **Colonic mucosa with dense inflammation consisting of lymphocytes, histiocytes, neutrophils, and scattered eosinophils.** |
|  |  |  | *2014<br>UC history | 0 | 0 | 0 | 0 | 0 | 0 | 0 | Chronic colonic mucosal injury. Fibrin-appearing eosinophilic amorphous deposition in the lamina propria capillaries. |
|  |  |  | 2018<br>UC history | 1 | 0 | 0 | 0 | 0 | 0 | 0 | No evidence of MC, collagenous colitis, or IBD |
| 13 | 2002 | 2019 | **\*2013**<br>**Fecal**<br>**incontinence** | **0** | **1** | **1** | **1** | **1** | **0** | **1** | **Colonic mucosa with acute cryptitis, glandular atrophy and architectural distortion in background of organizing fibrosis. Findings are suggestive of a long standing/chronic, possibly resolving injury. IBD appears less likely given lack of significant lymphoplasmacytic infiltrate.** |
|  |  |  | *2021<br>UC history | 1 | 0 | 0 | 0 | n/a (only polyps biopsied) | n/a | n/a |  |

\* denotes biopsy specimens that were available for direct evaluation by our gastrointestinal pathologist
Bold = on PPS, not bold = off PPS
CD = Crohn's disease
E = eosinophils
IBD = inflammatory bowel disease
MC = microscopic colitis
N = neutrophils
UC = ulcerative colitis

**Figure 2.**
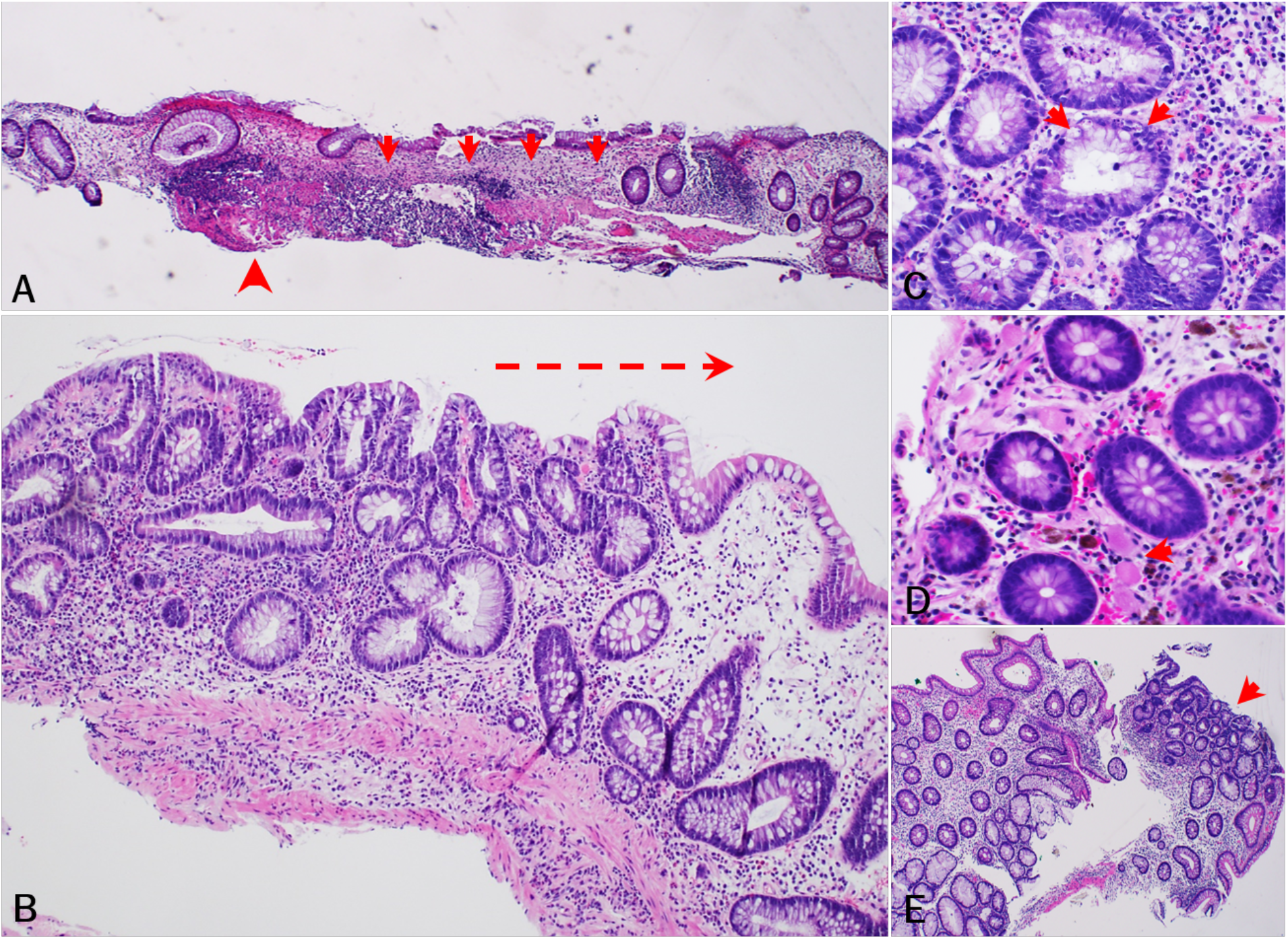
Histopathologic findings of PPS-related mucosal injury of colon (cohort study). A. [subject 5, 2015] Chronic mucosal injury characterized by apparent crypt dropout (arrow) and architectural disarray, patchy lamina propria fibrosis, and muscularis mucosa thickening (arrowhead). B. [subject 11, 2016] Injured colonic mucosa showing crypt architectural distortion and increased lamina propria lymphoplasmacytic inflammation with increased eosinophils and scattered neutrophils. Note lymphoplasmacytic inflammation progressing from left to right (arrow). C. [subject 4, 2018] Increased crypt epithelial apoptotic bodies. D. [subject 8, 2013] Fibrin-appearing eosinophilic amorphous deposition in lamina propria capillaries (arrow) and/or some stromal cells. Note melanosis coli also present. E. [subject 11, 2015] Low grade dysplasia (arrow) in non-targeted colonic biopsy. Note background colonic mucosa exhibiting crypt architecture abnormalities and regenerative changes.

All 11 participants reported their IBD or IBS symptoms have since improved. Seven of the nine with IBD received medical therapy. Symptoms improved with mesalamine (n=5), balsalazide (n=1), and budesonide (n=1). Three participants, who had all been using mesalamine, have since discontinued the medication and have not started alternative medications. One participant (subject 11) stopped using mesalamine following a colectomy, one (subject 2) stopped seven months after discontinuing PPS, and one (subject 8) stopped while still on PPS therapy.

Four participants (subjects 2, 3, 8, 12) had serial colonic biopsy specimens, including while on PPS and after stopping PPS therapy (Table 3). While using PPS, two (subjects 3, 12) of these four had rectosigmoid ulcerations on endoscopy, both of whose colonic lesions resolved following cessation of PPS use. Histopathologic signs of active colitis were observed for three participants (subjects 3, 8, 12) on PPS, none of which were evident in the biopsy specimens obtained when they were off therapy. All three managed their GI symptoms with mesalamine. Two participants (subjects 3,12) on PPS had histopathologic evidence of lamina propria lymphoplasmacytic inflammation, which was no longer present when they were off therapy. One participant (subject 2) on PPS had findings of chronic mucosal injury, which were no longer present when they were off PPS.

Two participants underwent a colectomy for colitis-associated dysplasia (Table 2). One participant (subject 11) underwent a colectomy for dysplasia one year after being diagnosed with UC, which was 15 years after starting PPS. This participant was on PPS at the time of the colectomy and had an approximate 2.42 kg cumulative exposure at that time. Another participant (subject 2) with a 0.84 kg cumulative PPS exposure had a partial colectomy for dysplasia in the context of microscopic colitis. This participant was initially diagnosed with UC 4.9 years into treatment with PPS but had been off PPS for 3.2 years prior to the colectomy. A third participant (subject 3) with a 2.18 kg cumulative PPS exposure is being monitored for a dysplastic lesion that may ultimately require partial colectomy.

Family history of IBD was present in one participant, IBS in none, and colorectal cancer in three. Exposure to other IC treatments included hydroxyzine (n=4), amitriptyline (n=4), gabapentin (n=3), cyclobenzaprine (n=2), methenamine (n=2), and phenazopyridine (n=1), none of which have been implicated in drug-induced colitis. Some of the 11 participants reported chronic use of medications that are associated with colitis, including statins (n=7), mesalamine (n=5), and NSAIDs (n=1).^14^

### Cross-Sectional Study

During the study period, 219 patients with IC were seen at the Emory Eye Center. Mean age was 60.8 (standard deviation, 15.1) years and 195 (89.0%) subjects were female. PPS exposure was documented in 80 (36.5%) subjects and was a statistically significant predictor of both the IBD [adjusted OR=3.3 (95% confidence interval, 1.2–8.8, p=0.02)] and the composite IBD+IBS [adjusted OR=3.3 (95% confidence interval, 1.5–7.3, p=0.002)] outcomes (Table 4). No other medication exposure was associated with an increased risk of either the IBD or IBD+IBS outcomes.

**Table 4.**
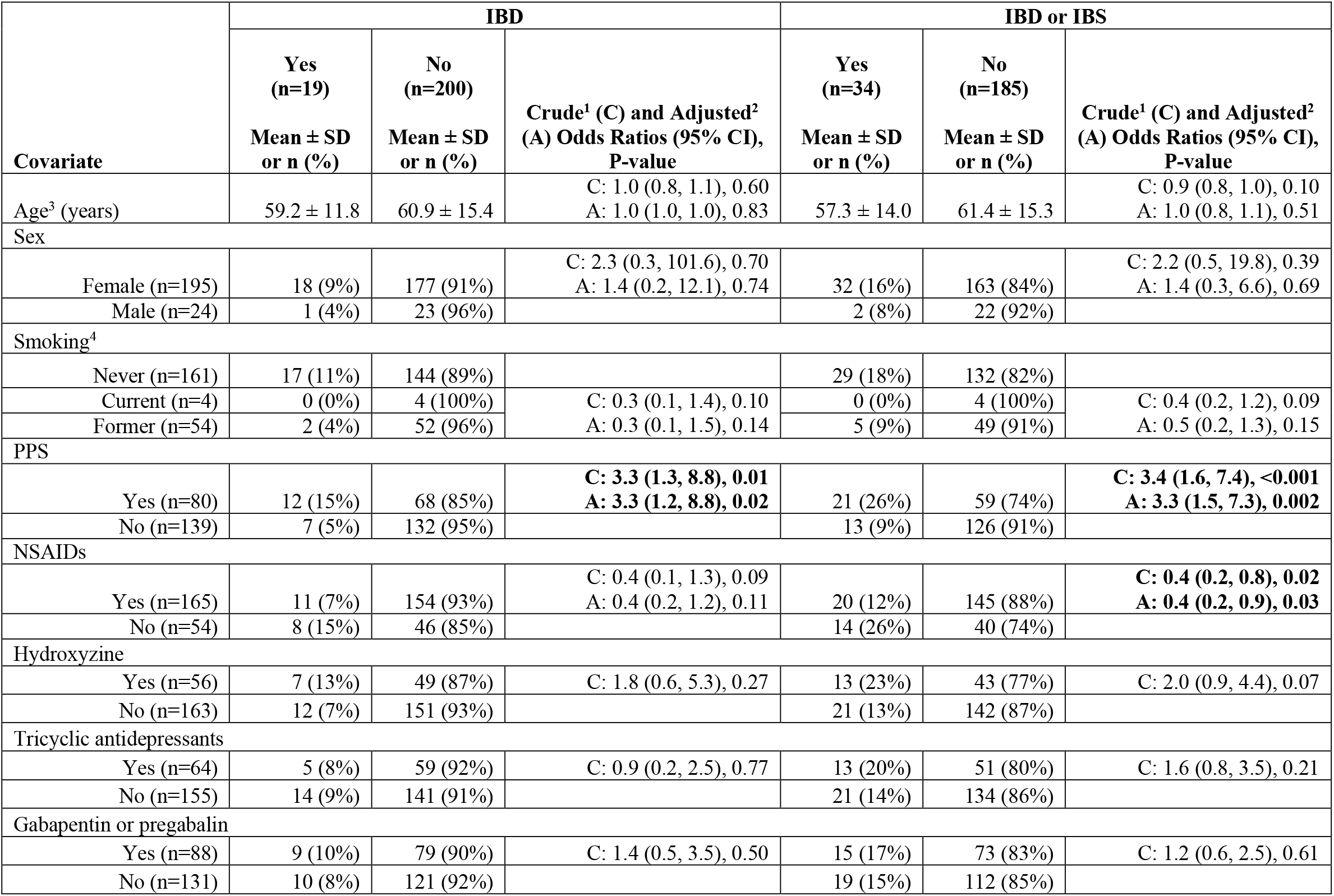
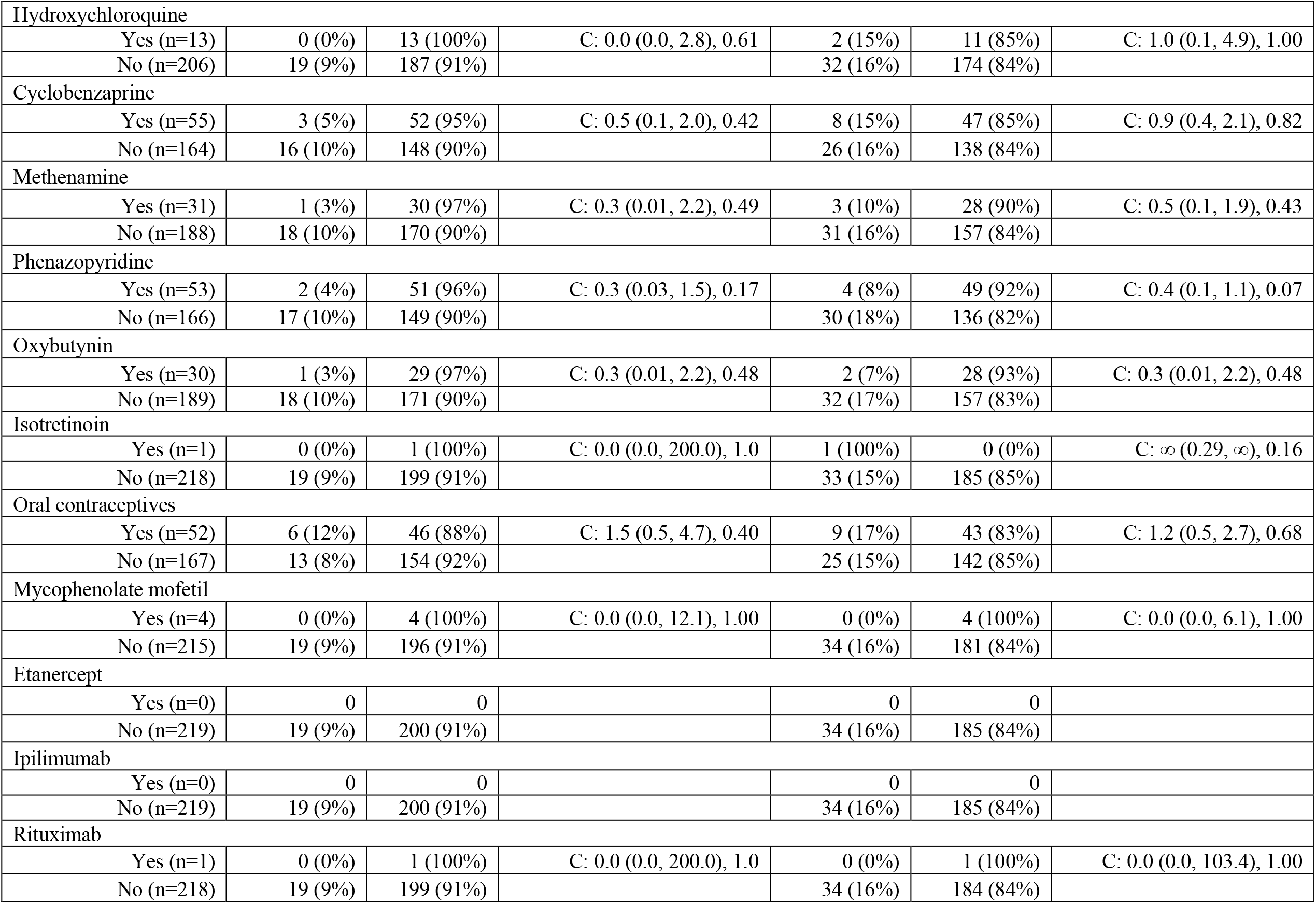
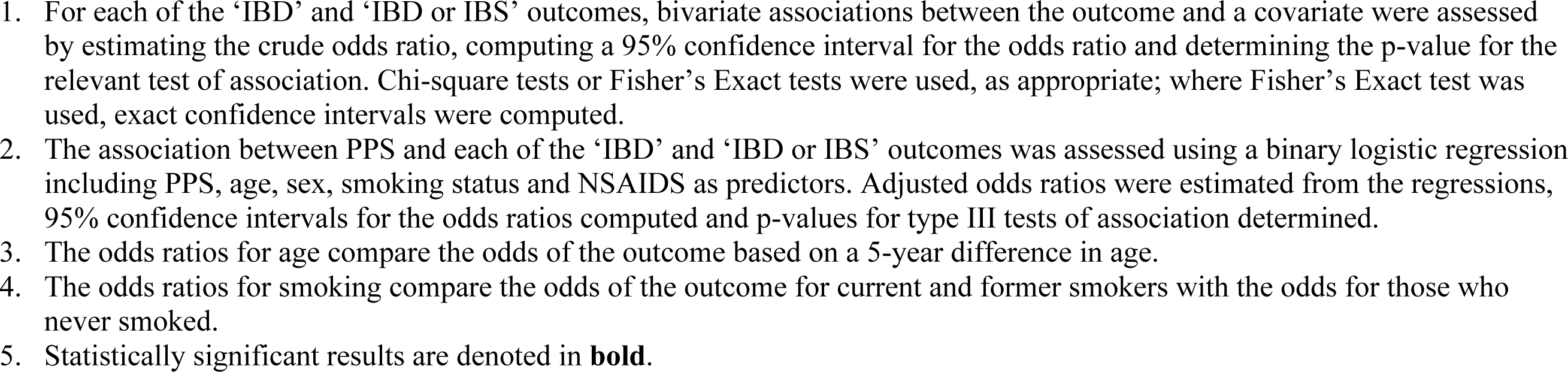
Study subject demographics and exposures to potential risk factors according to presence or absence of IBD, and IBD or IBS (cross-sectional study).

## Discussion

Our study demonstrates a novel association between exposure to PPS, a drug that has been widely used to manage IC for decades, and a possible drug-induced colopathy causing gastrointestinal symptoms. Among the 13 participants with high cumulative PPS exposure in our cohort study, 11 had received a clinical diagnosis of IBD or IBS after initiating PPS therapy. Our cross-sectional study of 219 subjects with IC corroborates this association, demonstrating that PPS was the only significant predictor of an IBD diagnosis among all IC therapies evaluated, with an adjusted OR of 3.3.

Similar to our findings with PPS maculopathy, this PPS-associated colopathy appears to manifest after long-term use of the drug, although further study is needed to establish a dose-response relationship. Symptoms ranging from diarrhea to pain and bleeding appeared to be moderate in severity but generally manageable with therapies such as mesalamine. Participants’ symptoms, need for treatment, and endoscopic and histopathologic findings all appeared to subside after PPS cessation. However, some patients were left with permanent sequelae after undergoing colectomy for colitis-associated dysplasia.

Histopathologically, seven of the ten participants with available histology slides exhibited chronic mucosal injury and five participants had findings of focal minimal to mild activity. No cases showed significant inflammation. Histopathological features of this PPS-associated colopathy included: 1) chronic mucosal injury characterized by crypt dropout and architectural disarray, and patchy lamina propria fibrosis and thickening of the muscularis mucosae, 2) patchy increased lamina propria lymphoplasmacytic inflammation with increased eosinophils, 3) increased crypt epithelial apoptotic bodies, 4) fibrin-appearing eosinophilic amorphous deposition in lamina propria capillaries and/or some stromal cells, and 5) foamy change of crypt epithelial cytoplasm and nonspecific superficial granulomas. These features are often observed in drug-induced colonic injury or colopathy.^15,16^ In some biopsies, low-grade dysplasia was also observed in non-polyp cases (non-targeted colonic biopsies), with the background colonic mucosa exhibiting only mild crypt architecture irregularity and regenerative changes. However, classic diagnostic features of IBD, such as basal crypt branching and diffuse basal plasmacytosis with lifting of the base of the crypts from the muscularis mucosae, were not evident in any biopsies.

Although the pathobiology of a potential PPS-induced colopathy is unclear, it is worth noting that PPS is similar in chemical structure to dextran sodium sulfate (Figure 1), which is widely used in a chemically-induced model of colitis in mice.^11^ PPS contains repeating sulfated xylan residues (2 sulfate groups per xylose residue), with a molecular weight range of 4,000-7,000 Daltons. Similarly, DSS is a highly sulfated polysaccharide (∼2.3 sulfates per glucosyl residue), with a molecular weight range of 5,000-14,000 Daltons. Due to the high degree of sulfation, both compounds have a high negative charge that may account for their biologic activities. Notably, differences in sulfation patterning, three-dimensional structure, and chain length may alter the pharmacological effect of each drug. Further, it is unclear whether PPS itself or a metabolite may be associated with both the retinal and gastrointestinal diseases.

Regarding the DSS colitis model, the earliest histologic changes are focal and not associated with inflammation.^17^ Mice that received DSS for only seven days developed an active chronic colitis histologically characterized by areas of both activity and inactivity, crypt loss and distortion, epithelial proliferation, and possibly even dysplasia.^17^ The earliest histologic features that were observed in the DSS colitis model, such as crypt loss without accompanying inflammation, are similar to what was seen in our study.

Although this is the first study to provide a detailed exploration of the association between PPS use and new-onset colopathy, prior studies have suggested an increased incidence of lower GI symptoms among PPS users. Indeed, current FDA labeling of the drug includes GI-related findings from an unblinded trial of 2499 IC subjects treated with PPS for varying durations.^4^ “ Colitis” is mentioned as one of 38 adverse events, at a frequency of ≤1%. The label cautions that patients with “ gastrointestinal ulcerations, polyps, or diverticula” should be “ carefully evaluated prior to starting ELMIRON.” ^4^ In the FDA medical review for PPS, it was noted that future phase IV studies would further explore the question of whether the GI side effects seen among the study subjects were due to the drug or to underlying diseases.^5^ A phase IV study (NCT00086684) was initiated in September 2003 and 369 subjects were assigned to one of three intervention arms for 24 weeks (PPS 100 mg once a day, PPS 100 mg three times a day, and placebo). Citing the lack of safety concerns raised on interim analysis, the phase IV study was terminated in June 2011.

Other studies have also reported on adverse GI effects among PPS users. In a phase I study investigating PPS for advanced cancers, 20 of 21 subjects experienced grade 1 or 2 gastrointestinal bleeding.^18^ Proctitis was the second most common toxicity, with nonspecific signs of inflammation on histology. Of note, in this study of subjects with multiple medical comorbidities, the use of concomitant medications, their potentially toxic effects, and potential drug interaction effects were not fully investigated. Another clinical study found that 4% of subjects receiving PPS 300 mg daily for 32 weeks experienced rectal hemorrhages.^19^ Finally, the Anderson et al. retrospective case series demonstrated high rates of colonic dysplasia among IBD patients who took PPS.^13^ Notably, a majority of these cases had an IBD diagnosis before PPS initiation, and this study did not specifically evaluate the association between PPS use and new IBD diagnoses.

Importantly, our findings do not establish a causal relationship between PPS and colopathy. An alternative explanation is that the underlying diagnosis of IC or another IC therapy is responsible for the GI disease. Indeed, multiple studies have demonstrated a link between IC and IBD. Perhaps the participants in our cohort study, all of whom used PPS for many years, had relatively severe underlying IC with more extensive involvement of other organ systems such as the colon. However, the strong association demonstrated here with two distinct studies, suggestion of temporality, and plausibility shown through studies of DSS are concerning. Further, prior studies of IC yield a 2% IBD prevalence rate that is much lower than the 85% colopathy rate in our cohort study. Finally, our cross-sectional study did not show evidence of confounding by any of the other IC therapies; PPS was the only drug with a statistically significant association with an IBD or IBS diagnosis.

Our cohort study was limited by use of retrospective data. All participants were off PPS prior to the baseline study visit. Most colonoscopy procedures were performed prior to consideration of a possible medication side effect, and biopsies of the background colonic mucosa were not consistently available. Thus, we were unable to directly compare endoscopic and histologic features from time points preceding PPS use, during PPS therapy, and following cessation of the drug in every subject. Despite these limitations, the clinicopathologic findings observed do provide a baseline for future investigations.

In conclusion, our study is suggestive of a novel association between PPS exposure and colopathy. These findings have important public health implications, given that PPS has been widely prescribed for decades. Although this may be a reversible condition, there is a significant burden of disease that often prompts invasive diagnostic procedures and treatments, and colonic dysplasia in some participants suggests that more deleterious outcomes are possible. Further study is warranted to investigate a potential causal relationship, evaluate the clinical impact of this condition, and explore the pathobiology of the disease.

## Data Availability

All data produced in the present work are contained in the manuscript.

